# Assessment of impending pancreatic cancer in a cohort of new onset diabetes on basis of biomarker trajectory

**DOI:** 10.64898/2026.08.06.26359908

**Authors:** Ehsan Irajizad, Camden Lopez, Suresh Chari, Jody Vykoukal, Rachelle Spencer, Yaxi Li, Jennifer B. Dennison, Eugene J. Koay, Florencia McAllister, Michael Kim, Matthew Young, Phil A. Hart, Willam Fisher, Stephen K. Vandeneeden, Bechien U. Wu, Ziding Feng, Samir Hanash, Anirban Maitra, Johannes F. Fahrmann, the Consortium for the Study of Chronic Pancreatitis, Diabetes, and Pancreatic Cancer (CPDPC)

**Affiliations:** Department of Biostatistics, The University of Texas MD Anderson Cancer Center, Houston, TX; Biostatistics Program, Public Health Sciences Division, Fred Hutchinson Cancer Research Center, Seattle, Washington, USA; Department of Clinical Cancer Prevention, The University of Texas MD Anderson Cancer Center, Houston, TX; Department of Gastroenterology Hepatology and Nutrition, The University of Texas MD Anderson Cancer Center, Houston, TX; Department of GI Radiation Oncology, The University of Texas MD Anderson Cancer Center, Houston, TX; Department of Genetics, The University of Texas MD Anderson Cancer Center, Houston, TX; Department of Gastrointestinal Medical Oncology, The University of Texas MD Anderson Cancer Center, Houston, TX; Department of Surgical Oncology, The University of Texas MD Anderson Cancer Center, Houston, TX; National Cancer Institute, Division of Cancer Prevention, National Cancer Institute, Bethesda, MD, USA; Division of Gastroenterology, Hepatology, and Nutrition, The Ohio State University Wexner Medical Center, Columbus, OH, USA; Department of Surgery, Baylor College of Medicine, Houston, Texas, 77054; Division of Research, Kaiser Permanente Northern California, Oakland, CA, United States; Center for Pancreatic Care, Southern California Permanente Medical Group, Department of Gastroenterology, Kaiser Permanente Los Angeles Medical Center, Los Angeles, California, USA; Department of Pathology and Medicine, Perlmutter Cancer Center, NYU Grossman School of Medicine, New York, NY

## Abstract

**PURPOSE:** To assess the predictive performance of panel protein biomarkers as well as an established algorithm that considers repeat biomarker testing for risk prediction of PDAC among a prospective cohort of patients with New-onset diabetes.

**PATIENTS AND METHODS:** A panel of protein biomarkers (CA19-9, CA125, CEA, LRG1, REG3A and TIMP1) were assayed in 6,516 serially collected pre-diagnostic plasma samples from 2,121 NOD patients from the Consortium of Chronic Pancreatitis Diabetes and Pancreatic Cancer (CPDPC)-initiated NOD study who completed the 3-year study follow-up period. The specimen set included 25 pre-diagnostic samples from the 12 PDAC cases diagnosed during study follow-up. We applied a single threshold (ST) method, which considers biomarker levels at a single time point, as well as a previously established parametrical empirical Bayes (PEB) algorithm, which considers prior biomarker measurements, with ‘case’ calls made based on pre-specified cutoffs corresponding to 1% 1-year risk. Resultant biomarker data as well as case calls were provided to the EDRN Data Management and Coordinating Center as part of a Prospective-sample-collection-Retrospective-Blinded-Evaluation (ProBE)-compliant Phase 3 biomarker validation study. Area under the Receiver Operating Characteristic Curves (AUC), sensitivity, specificity, population-level positive predictive value (PPV), and negative predictive value (NPV) are reported.

**RESULTS:** The 3-year incidence of PDAC in the NOD cohort was 0.57%. When considering PDAC vs non-cancer controls, respective AUCs of individual protein biomarkers ranged from 0.52-0.94, with CA19-9 achieving the highest overall performance of 0.94 (95% CI: 0.86-1.00). At the pre-defined 1% 1-year risk threshold, CA19-9 yielded sensitivity of 83.3% at 97.2% specificity. Additional markers CEA, CA125, and TIMP1 demonstrated sensitivity of 33.3%, 41.7%, and 8.3%, respectively. In a subset of patients, CA19-9 first tested ‘positive’ at a median (interquartile range [IQR]) of 7 months (4 to 14 months) prior to clinical PDAC diagnosis. Of the two PDAC cases missed by CA19-9 using the ST method, one (diagnosed with stage III PDAC) was detected using the PEB^CA19-9^ algorithm.

**CONCLUSION:** In the setting of adult new onset diabetes, CA19-9 is a readily available and promising biomarker that can be leveraged for earlier detection of an underlying pancreatic cancer. Additional protein biomarkers may improve sensitivity for earlier detection of PDAC among cases with low CA19-9.

## Introduction

Pancreatic ductal adenocarcinoma (PDAC) continues to be among the leading cause of cancer-related mortality in the world, despite representing 3% of all cancer cases.^1^ Dismal overall 5-year survival rates of less than 13% are attributed to most (80-85%) individuals presenting with advanced stage disease, when curative-intent surgical resection is no longer possible.^2-5^ Population-based screening for PDAC is currently not feasible due to low prevalence of the disease among otherwise average-risk individuals and potential harms associated with false-positive results. Instead, focus has shifted towards sub-groups of individuals who are at elevated risk of PDAC and who may benefit from surveillance.^6,7^

Patients ≥50 years of age with new onset diabetes (NOD) have received considerable attention as an elevated risk population. To-date, several studies have reported ∼6-8 fold higher risk of PDAC among NOD patients compared to those in the general population, with the highest risk occurring within 3 years of diabetes onset.^8-14^ Although patients with NOD are at higher risk compared to that of the general population, it is widely recognized that further stratification is needed to better select for the highest-risk subgroup of NOD patients for targeted screening.^10^

Recently, we demonstrated lead-time utility of CA19-9 as well as other pertinent PDAC-associated protein biomarkers, including CEA, CA125, LRG1, REG3A, and TIMP1, for earlier detection of PDAC in the Prostate Lung Colorectal and Ovarian (PLCO) cohort. We reported an exponential rise in CA19-9 starting 2 years prior to a clinical PDAC diagnosis, reaching sensitivity of 50% at >99% specificity for early-stage disease considering samples collected within 6 months of diagnosis.^15,16^ Using an independent set of PLCO samples, we further demonstrated that an adaptive algorithm that considers repeat CA19-9 testing improves sensitivity and results in an earlier ‘positive’ signal by ∼222 days (7.4 months) compared to a single timepoint measurement. Notably, in this study, a PEB algorithm based on repeat measurements of TIMP1 yielded an incremental improvement in sensitivity of 14 % without loss of specificity for identifying PDAC cases with low CA19-9 levels. ^15,16^

In the current study, we performed a Prospective-sample-collection-Retrospective-Blinded-Evaluation (PRoBE)-compliant Early Detection Research Network (EDRN)-defined Phase 3 biomarker validation study to test the merits of CA19-9 for risk prediction of PDAC among 2,120 NOD patients that were prospectively followed for 3 years. The complementarity of CEA, CA125, LRG1, REG3A, and TIMP1 were also evaluated. We report sensitivity and specificity as well as positive and negative predictive values when considering a single timepoint measurement and when considering algorithm that incorporates the longitudinal trajectory of protein biomarkers.

### Methods NOD Cohort

Detailed information regarding the NOD cohort^17^ is provided in the **Supplemental Methods**. The current study leveraged all available baseline and serially collected (6, 12, and 24 months) plasma samples (N= 6,516) from the 2,121 NOD participants that completed the 3-year study follow-up period. Patient characteristics are described in **Table 1**. Of the 2,121 NOD patients, 12 were diagnosed with PDAC (**Table 1** and **Supplemental Tables S1-2**) and 125 were diagnosed with other non-PDAC cancers during course of study follow-up.

**Table 1.**
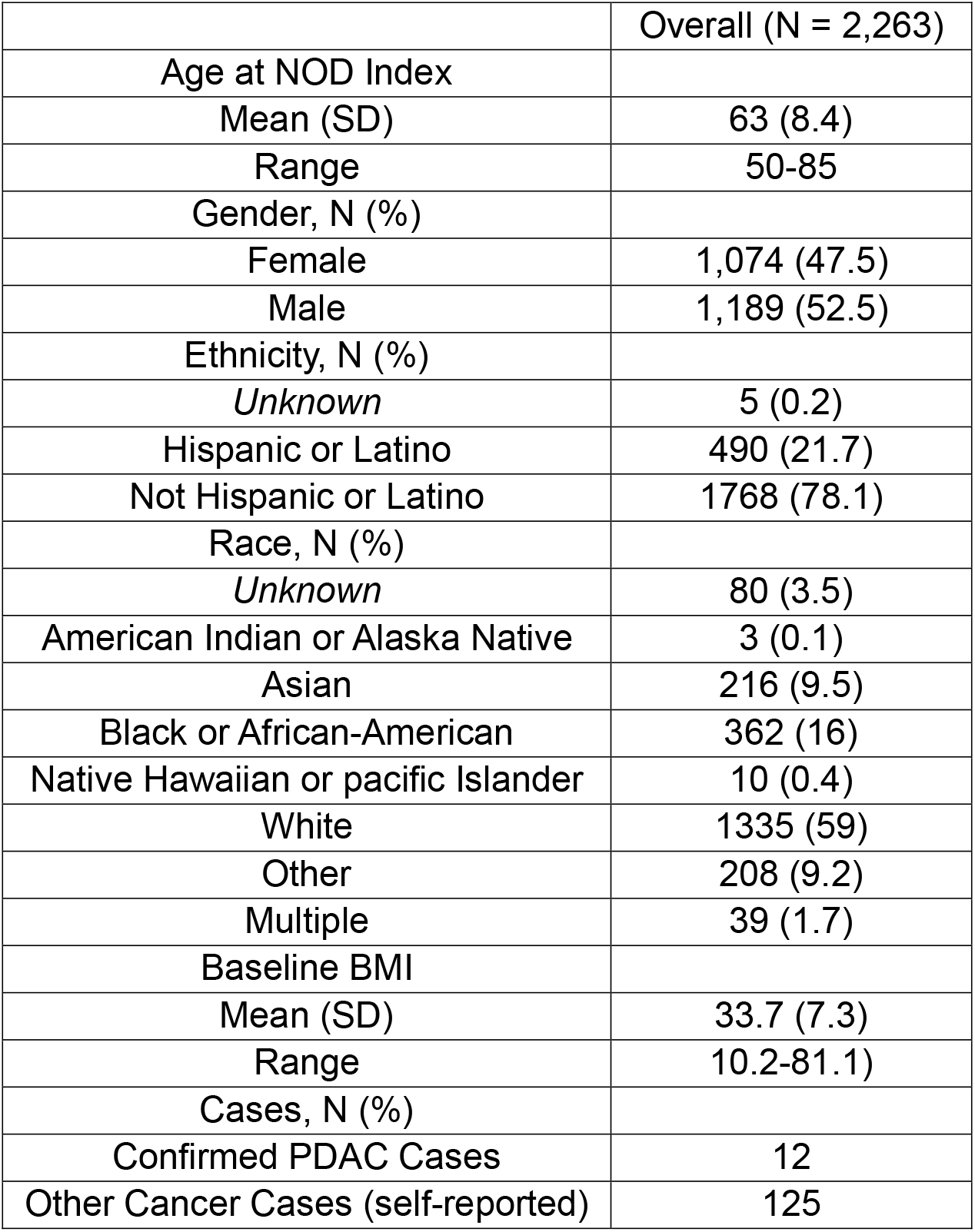
Patient and tumor characteristics for the NOD cohort.

### Enzyme-linked Immunosorbent Assays

Plasma protein concentrations for CA19-9, CEA, CA125, LRG1, REG3A, and TIMP1 were determined by bead-based ELISA assays using Luminex multiplexed assay technology ([CA19-9] HCCBP1–58MAG, [LRG1] HCVD6MAG-67K, [TIMP1] HTMP1MAG-54K; Millipore, Bedford, MA), [REG3A] CUST0I704-BULK, and [CEA and CA125] HCCBP1MAG-58K, as previously described.^15,16^

### Statistical Analyses

We applied a single threshold (ST) method, which considers the value of the biomarker at a single time point, as well as a parametrical empirical Bayes (PEB) algorithm, which considers all existing measurements of the biomarker and adjusts the biomarker threshold to reflect the participant’s biomarker history.^16^ For the PEB algorithm, we used fixed model tuning parameters from our prior publication. No re-training was performed.

Individual CA19-9, CEA, CA125, LRG1, REG3A, and TIMP1 readouts as well as ‘case’ calls based on pre-specified cutoffs corresponding to 1% 1-year risk based on either the single-time point or PEB method were provided to the EDRN Data Management and Coordinating Center for statistical analyses.

Area under the Receiver Operating Characteristic curves (AUC), sensitivity, specificity, positive predictive values (PPV) and negative predictive values (NPV) were calculated at the specimen-level and at the patient-level (i.e. a patient is classified as a ‘case’ if any of their serial samples show a biomarker value exceeding the predefined threshold). The 95% confidence intervals (CI) for AUCs were estimated using the DeLong method.

## Results

Of the 2,121 NOD patients that completed the 3-year follow-up study period, 12 were diagnosed with PDAC corresponding to a 3-year risk of 0.57% (**Table 1**), which is consistent with 3-year risk estimates reported by others.^10,11,14,18^ Of the 12 PDAC diagnoses, 6 (50%) were diagnosed within 1 year, 3 (25%) within 1-2 years, and 3 (25%) within 2-3 years of study enrollment. The majority of PDAC cases were clinically diagnosed with advanced (III-IV) stage (10 out of 12) (**Supplemental Tables S1-2**).

Receiver operating characteristic curves for CA19-9, CEA, CA125, LRG1, REG3A, and TIMP1 in distinguishing PDAC cases from non-cancer controls, other cancer types, as well as non-PDAC cases (non-cancer controls + other cancer types) at the patient level are provided in **Supplemental Table S3**. When considering PDAC vs non-cancer controls, respective AUCs of individual protein biomarkers ranged from 0.52-0.94, with CA19-9 achieving the highest overall performance of 0.94 (95% CI: 0.86-1.00) (**Supplemental Table S3**).

At the 1% 1-year risk threshold, CA19-9 yielded a sensitivity of 83.3% and specificity of 97.2% for risk prediction of PDAC in comparison to non-cancer controls (**Figure 1, Table 2; Supplemental Tables S4-6**). Notably, 7 (5.6%) of the 125 other non-PDAC cancers were detected by CA19-9 (**Table 2; Supplemental Table S5**). Although no marker outperformed CA19-9, additional markers CEA, CA125, and TIMP1 demonstrated sensitivity of 33.3%, 41.7%, and 8.3%, respectively, at the pre-defined 1% 1-year risk thresholds (**Supplemental Table S4**).

**Table 2.**
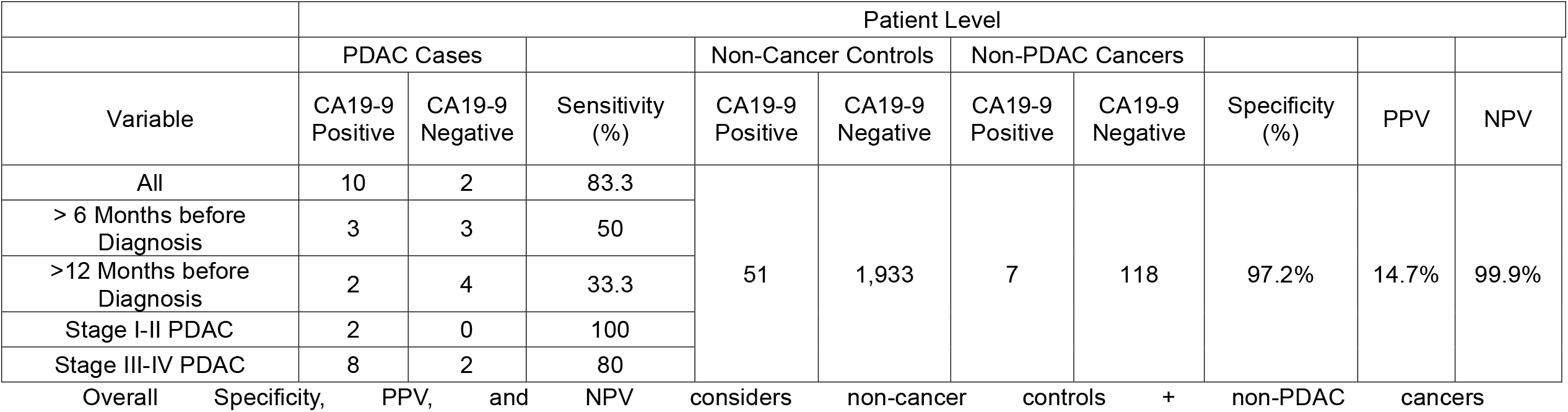
Performance estimates of single-timepoint CA19-9 at the patient-level for detection of PDAC in the NOD Cohort.

**Figure 1.**
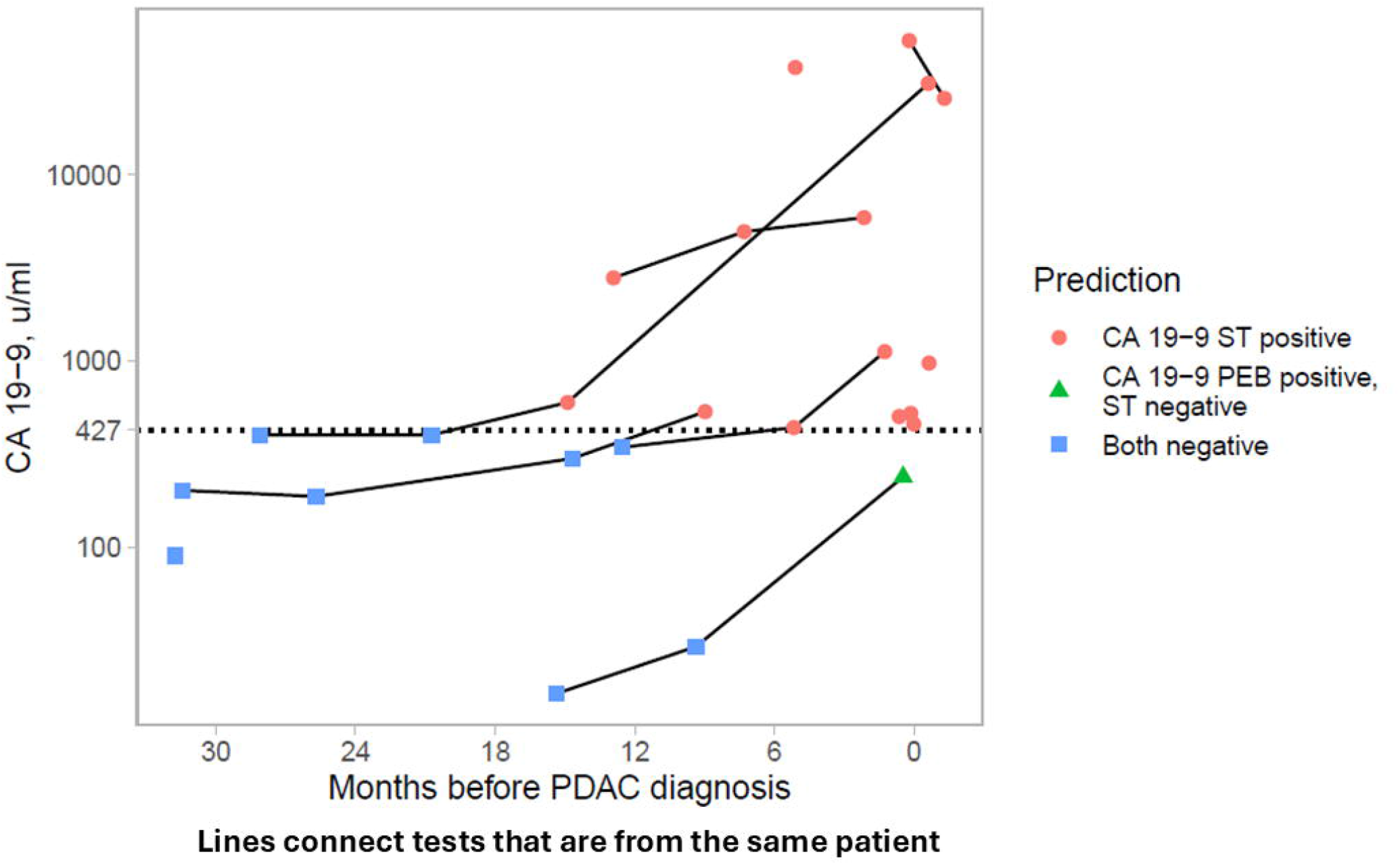
Test positivity for CA19-9 at the specimen level for the 12 PDAC cases in the NOD Cohort. Orange circle-positive for CA19-9 based on single timepoint (ST) measurement; green circle-Positive based on PEB^CA19-9^. Blue Circle-negative for CA19-9 for both ST^CA19-9^ or PEB^CA19-9^. Additional information is provided in **Supplemental Table S2**.

At the specimen level, CA19-9 first tested ‘positive’ (i.e. met or exceeded the 1% 1-year risk threshold) in a sub-set of cases (N=6) at a median (interquartile range [IQR]) of 7 months (4 to 14 months) prior to clinical PDAC diagnosis, with two of these cases testing positive for CA19-9 >12 months before a clinical diagnosis (**Figure 1; Supplemental Table S1**).

Notably, two cases were not identified by CA19-9 using a typical ST approach (**Figure 1**). Of these two cases, one (stage III) was test ‘positive’ based on the PEB^CA19-9^ algorithm (**Figure 1**). The other case had a single timepoint collected at >30 months before diagnosis (**Supplemental Table S1**).

## Discussion

A recent prospective, observational study of 18,838 adults _≥_50 years of age with NOD demonstrated a race-adjusted 3-year PDAC incidence of 0.62%^19^, consistent with the 3-year PDAC incidence of 0.57% observed in the NOD cohort. Despite higher risk, screening for PDAC with any test among NOD patients is not recommended as the incidence among this population is still too low to justify screening. The goal of our study was to determine whether CA19-9, a routinely used laboratory test, would identify individuals with NOD who are at sufficiently elevated risk of having PDAC to now warrant additional image-based screening. We focused on a CA19-9 threshold corresponding to _≥_1% 1-year risk. At this risk threshold, CA19-9 achieved adequate high sensitivity (>80%) while maintaining high specificity. Importantly, in a subset of patients, CA19-9 first tested ‘positive’ at a median of 9 months prior to clinical PDAC diagnosis. On the basis of these findings, we anticipate that testing of CA19-9 in NOD patients would result in a shift towards earlier detection of resectable disease, and that risk-based PDAC screening in patients with NOD is cost-effective even if a modest (>25%) shift towards resectable disease is achieved.^20^

### Strengths and Limitations

There are several strengths to our study. Inclusion of other cancer types prevalent among NOD patients additionally allows for assessment of the specificity of CA19-9 for PDAC compared to other cancer types. Another strength of the NOD cohort is a high representation of non-White individuals, allowing for broader generalizability of findings. Limitations to our study include the small number of PDACs, assaying of CA19-9 with Luminex non-IVD assay rather than clinical CA19-9 testing. The frequency of patients with fructosyltransferase deficiency is unknown in the NOD Cohort. It is plausible that ‘non-case’ participants may go on to developed PDAC beyond the 3-year NOD study period.

In summary, our findings support the potential to use of CA19-9 as part of routine testing for NOD patients to inform on the need for close monitoring and screening for earlier detection of disease. A larger prospective study is warranted to get more precise and definitive performance measures of CA19-9 on pre-diagnostic blood in NOD patients. Additional protein biomarkers, such as CEA, CA125, and TIMP1, may provide incremental improvement in sensitivity.

## Data Availability

This study did not generate new reagents.
There are restrictions to the availability of human biospecimens due to existing MTA.
Relevant data supporting the findings of this study are available within the Article and Supplemental Materials.
No new code was generated for this study.
Any additional information required to reanalyze the data reported in this paper is available from the Lead Contact upon request.

## Acknowledgements

This project was Supported in part through the generous philanthropic contributions to The University of Texas MD Anderson Cancer Center Moon Shots Program, Pancreatic Cancer North America and the Warren Y. Soper Charitable Trust, NCI U01CA200468, and the Sheikh Khalifa bin Zayed Foundation. Research reported in this study was supported by National Cancer Institute and National Institute of Diabetes and Digestive and Kidney Diseases of the National Institutes of Health under award numbers: 2 U01 DK108314, 1 U01 DK126365, 2 U01 DK108306, 2 U01 DK108323, 1 U01 DK126300, 2 U01 DK108326, 2 U01 DK108334, 2 U01 DK108327, 2 U01 DK108288, 2 U01 DK108328, 2 U01 DK108300, and 2 U01 DK108320.”*

## Author Contributions

### Conception and Design

Ehsan Irajizad, Suresh Chari, Matthew Young, Phil A Hart, William Fisher, Stephen K. Vandeneeden, Bechien Wu, Ziding Feng, Anirban Maitra, Samir Hanash, and Johannes F. Fahrmann.

### Collection and assembly of data

Ehsan Irajizad, Jody Vykoukal, Rachelle Spencer, Yaxi Li, and Johannes F. Fahrmann.

### Data analysis and interpretation

Ehsan Irajizad, Camden Lopez, Ziding Feng, and Johannes F. Fahrmann.

### Writing of original manuscript

Ehsan Irajizad, and Johannes F. Fahrmann.

### Editing of manuscript

Camden Lopez, Suresh Char, Jody Vykoukal, Rachelle Spencer, Yaxi Li, Jennifer B. Dennison, Eugene J. Koay, Florencia McAllister, Michael Kim, Joann Rinaudo, Phil A. Hart, William Fisher, Stephen K. Vandeneeden, Bechien Wu, Ziding Feng, Samir Hanash, and Anirban Maitra.

## Final approval of manuscript

All authors

## Accountable for all aspects of the work

All authors

