## Supplemental Materials for "Assessment of impending pancreatic cancer in a cohort of new onset diabetes on basis of biomarker trajectory"

**New Onset Diabetes (NOD) Cohort**

In 2015, the National Cancer Institute and the National Institute for Diabetes and Digestive and Kidney Diseases initiated the Consortium for the study of Chronic Pancreatitis, Diabetes, and Pancreatic Cancer (CPDPC).^1^ One of the objectives of the CPDPC was to assemble a cohort of 10,000 subjects ≥ 50 years of age with new-onset diabetes (NOD) from 11 participating study centers, with each subject participating for up to 3 years from the date they met criteria for NOD (Clinical Trial NCT03731637) under IRB approved protocols 17-011305, U01DK108328, A211701, NCI-2018-01307, and RG1001811. Detailed study definitions and criteria for the NOD study are discussed elsewhere.^2^

The primary objective of the NOD study was to i) estimate of the 3-year probability of PDAC in NOD; ii) establish a biorepository of clinically annotated bio-specimens, from pre-symptomatic PDAC and non-case control new-onset type 2 diabetes mellitus subjects. All enrolled subjects were followed for collection of clinical data and bio-specimen samples at baseline (i.e., at the time of recruitment), and subsequently at 6-, 12-, and 24-months post enrollment. iii) to use biospecimens for Early Detection Research Network (EDRN) defined- Phase 3 validation studies of promising biomarkers for earlier detection of incidence PDAC in NOD patients; and iv) to provide a platform for development of future interventional screening protocols for early detection of PDAC in NOD patients that also incorporates imaging studies and clinical algorithms.^2^

The current study leveraged all available baseline and serially collected (6, 12, and 24 months) plasma samples (N= 6,516) from the 2,121 NOD participants that completed the 3-year study follow-up period. Of the 2,121 NOD patients, 12 (0.57%) were diagnosed with PDAC and 125 (5.9%) were diagnosed with other non-PDAC cancers.

**Supplemental Table S1. Patient and tumor characteristics for the NOD cohort.**

|  | Overall (N = 2,263) |
| --- | --- |
| Age at NOD Index |  |
| Mean (SD) | 63 (8.4) |
| Range | 50-85 |
| Gender, N (%) |  |
| Female | 1,074 (47.5) |
| Male | 1,189 (52.5) |
| Ethnicity, N (%) |  |
| *Unknown* | 5 (0.2) |
| Hispanic or Latino | 490 (21.7) |
| Not Hispanic or Latino | 1768 (78.1) |
| Race, N (%) |  |
| *Unknown* | 80 (3.5) |
| American Indian or Alaska Native | 3 (0.1) |
| Asian | 216 (9.5) |
| Black or African-American | 362 (16) |
| Native Hawaiian or pacific Islander | 10 (0.4) |
| White | 1335 (59) |
| Other | 208 (9.2) |
| Multiple | 39 (1.7) |
| Baseline BMI |  |
| Mean (SD) | 33.7 (7.3) |
| Range | 10.2-81.1) |
| Cases, N (%) |  |
| Confirmed PDAC Cases | 12 |
| Other Cancer Cases (self-reported) | 125 |

**Supplemental Table S2. Time from baseline entry to clinical diagnosis for the 12 PDAC cases in the NOD cohort.**

| Case | Sampling Timepoints  (M: Months)¥ | Diagnosis Time | Stage at Diagnosis |
| --- | --- | --- | --- |
| 1 | Baseline | (-)1 Month | IA |
| 2 | Baseline | Baseline | III |
| 3 | Baseline, 2M | Baseline | IV |
| 4 | Baseline | Baseline | IIB |
| 5 | Baseline, 6M, 11M | 13 Months | IV |
| 6 | Baseline | 1 Month | IV |
| 7 | Baseline | 5 Months | IV |
| 8 | Baseline, 7M, 11M | 13 Months | III |
| 9 | Baseline, 7M, 13M, 29M | 28 Months | IV |
| 10 | Baseline, 6M, 15M | 15 Months | III |
| 11 | Baseline, 6M, 17M, 22M | 31 Months | IV |
| 12 | Baseline | 32 Months | IV |

¥ number of serial timepoints available and time at which samples were collected following study enrollment

**Supplemental Table S3. Predictive performance of individual biomarkers for distinguishing PDAC from non-cancer controls, other cancer types, and non-PDAC cases (non-cancer controls + other cancer types) in the NOD Cohort.** Performance estimates are provided at the patient level.

|  | PDAC vs  Non-Cancer Controls | PDAC vs  Other Cancers | PDAC vs Non-PDAC Controls^ |
| --- | --- | --- | --- |
| Marker | AUC (95% CI) | AUC (95% CI) | AUC (95% CI) |
| CA19-9 | 0.94 (0.86-1.00) | 0.92 (0.83-1.00) | 0.94 (0.86-1.00) |
| CEA | 0.76 (0.60-0.93) | 0.74 (0.57-0.91) | 0.76 (0.60-0.93) |
| CA125 | 0.76 (0.58-0.94) | 0.72 (0.53-0.91) | 0.76 (0.58-0.94) |
| LRG1 | 0.52 (0.34-0.70) | 0.49 (0.29-0.68) | 0.52 (0.34-0.70) |
| REG3A | 0.58 (0.42-0.74) | 0.56 (0.40-0.73) | 0.58 (0.42-0.74) |
| TIMP1 | 0.58 (0.40-0.78) | 0.56 (0.37-0.75) | 0.58 (0.40-0.76) |

^ Non-Cancer Controls + Other Cancers

**Supplemental Table S4. Performance estimates of individual biomarkers based on pre-specified risk thresholds (1% 3-year risk) at the patient-level for detection of PDAC in the NOD Cohort.**

| N | 12 | 1984 | 125 |
| --- | --- | --- | --- |
| Marker | # Positive (%)  PDAC Cases | # Positive (%)  Non-Cancer Controls | # Positive (%)  Other Cancer Cases |
| CA19-9 | 10 (83) | 51 (3) | 7 (6) |
| CEA | 4 (33) | 48 (2) | 6 (5) |
| CA125 | 2 (17) | 64 (3) | 7 (6) |
| LRG1 | 0 (0) | 93 (5) | 6 (5) |
| REG3A | 0 (0) | 75 (4) | 3 (2) |
| TIMP1 | 1 (8) | 71 (4) | 5 (4) |

**Supplemental Table S5. Performance estimates of CA19-9 at the specimen-level for detection of PDAC in the NOD Cohort.**

|  | Specimen Level | | | | | | | | | | | |
| --- | --- | --- | --- | --- | --- | --- | --- | --- | --- | --- | --- | --- |
|  | PDAC Cases | |  | Non-Cancer Controls | | Non-PDAC Cancers | | |  | |  | |
| Variable | CA19-9 Positive | CA19-9  Negative | Sensitivity (%) | CA19-9 Positive | CA19-9  Negative | CA19-9 Positive | CA19-9  Negative | Specificity (%) | | PPV | | NPV |
| All | 15 | 10 | 60.0 | 88 | 6,033 | 11 | 359 | 98.5 | | 13.2 | | 99.8 |
| > 6 Months before Diagnosis | 4 | 9 | 30.8 |  |  |  |  |  |  |  |  |  |
| >12 Months before Diagnosis | 2 | 8 | 20.0 |  |  |  |  |  |  |  |  |  |
| Stage I-II PDAC | 2 | 0 | 100.0 |  |  |  |  |  |  |  |  |  |
| Stage III-IV PDAC | 13 | 10 | 56.5 |  |  |  |  |  |  |  |  |  |

Specificity, PPV, and NPV considers non-cancer controls + non-PDAC cancers

**Supplemental Table S6. Sensitivity of CA19-9 for detection PDAC cases at varying specificity levels based on non-cancer controls in the NOD Cohort.**

| Sample-level specificity | Sample-level sensitivity | Patient-Level Sensitivity |
| --- | --- | --- |
| 85% | 21/25 (84%) | 11/12 (91.7%) |
| 88% | 20/25 (80%) | 11/12 (91.7%) |
| 90% | 20/25 (80%) | 11/12 (91.7%) |
| 95% | 18/28 (72%) | 10/12 (83.3%) |
